# Proteogenomic integration reveals CXCL10 as a potentially downstream causal mediator for IL-6 signaling on atherosclerosis

**DOI:** 10.1101/2023.03.24.23287543

**Authors:** Savvina Prapiadou, Luka Živković, Barbara Thorand, Marc J. George, Sander W. van der Laan, Rainer Malik, Christian Herder, Wolfgang Koenig, Thor Ueland, Ola Kleveland, Pal Aukrust, Lars Gullestad, Jürgen Bernhagen, Gerard Pasterkamp, Annette Peters, Aroon D. Hingorani, Jonathan Rosand, Martin Dichgans, Christopher D. Anderson, Marios K. Georgakis

## Abstract

**Background:** Genetic and experimental studies support a causal involvement of interleukin-6 (IL-6) signaling in atheroprogression. While trials targeting IL-6 signaling are underway, any benefits must be balanced against an impaired host immune response. Dissecting the mechanisms that mediate the effects of IL-6 signaling on atherosclerosis could offer insights about novel drug targets with more specific effects.

**Methods:** Leveraging data from 522,681 individuals, we constructed a genetic instrument of 26 variants in the gene encoding the IL-6 receptor (IL-6R) that proxied for pharmacological IL-6R inhibition. Using Mendelian randomization (MR), we assessed its effects on 3,281 plasma proteins quantified with an aptamer-based assay in the INTERVAL cohort (n=3,301). Using mediation MR, we explored proteomic mediators of the effects of genetically proxied IL-6 signaling on coronary artery disease (CAD), large artery atherosclerotic stroke (LAAS), and peripheral artery disease (PAD). For significant mediators, we tested associations of their circulating levels with incident cardiovascular events in a population-based study (n=1,704) and explored the histological, transcriptomic, and cellular phenotypes correlated with their expression levels in samples from human atherosclerotic lesions.

**Results:** We found significant effects of genetically proxied IL-6 signaling on 70 circulating proteins involved in cytokine production/regulation and immune cell recruitment/differentiation, which correlated with the proteomic effects of pharmacological IL-6R inhibition in a clinical trial. Among the 70 significant proteins, genetically proxied circulating levels of CXCL10 were associated with risk of CAD, LAAS, and PAD with up to 67% of the effects of genetically downregulated IL-6 signaling on these endpoints mediated by decreases in CXCL10. Higher midlife circulating CXCL10 levels were associated with a larger number of cardiovascular events over 20 years, whereas higher *CXCL10* expression in human atherosclerotic lesions correlated with a larger lipid core and a transcriptomic profile reflecting immune cell infiltration, adaptive immune system activation, and cytokine signaling.

**Conclusions:** Integrating multiomics data, we found a proteomic signature of IL-6 signaling activation and mediators of its effects on cardiovascular disease. Our analyses suggest the interferon-γ-inducible chemokine CXCL10 to be a potentially causal mediator for atherosclerosis in three vascular compartments and as such could serve as a promising drug target for atheroprotection.

## INTRODUCTION

Anti-inflammatory treatments have emerged as a promising approach for lowering risk of atherosclerotic cardiovascular disease.^1^ The interleukin-6 (IL-6) pathway has attracted major attention due to converging evidence supporting its relevance in atherosclerosis.^2^ First, pharmacological inhibition of IL-6 or its receptor (IL-6R) leads to reductions in atherosclerotic lesion formation in mouse models of atherosclerosis.^3, 4^ Second, prospective cohort studies have consistently found circulating levels of IL-6 to be associated with manifestations of atherosclerotic disease including coronary artery disease^5^ and ischemic stroke.^6^ Third, human genetic studies have shown that polymorphisms in the gene encoding the IL-6 receptor (IL-6R) resulting in downregulated IL-6 signaling activity are associated with a lower lifetime risk of coronary artery disease,^7^ atherosclerotic ischemic stroke,^8^ peripheral artery disease,^9^ and abdominal aortic aneurysm,^10^ as well as a favorable cardiometabolic profile.^11^

In line with these results, the Canakinumab Anti-Inflammatory Thrombosis Outcomes Study (CANTOS) that tested a monoclonal antibody targeting IL-1β, which is an upstream regulator of IL-6 signaling, showed considerable reductions in major adverse cardiovascular events among patients with a recent history of myocardial infarction.^12^ Interestingly, *post hoc* analyses of CANTOS showed that canakinumab was particularly protective among patients achieving substantial decreases in IL-6 levels, thus providing indirect evidence that interfering with IL-6 signaling could reduce cardiovascular risk.^13^ On the basis of these findings, a recent phase 2 trial (RESCUE) tested ziltivekimab, a monoclonal antibody that is directed against IL-6, and found that it effectively and safely reduces biomarkers of inflammation and thrombosis among patients with chronic kidney disease and evidence of inflammation (high-sensitivity C-reactive protein (CRP) levels ≥ 2 mg/L).^14^ The cardiovascular benefit of this approach remains unknown as an ongoing large-scale phase 3 cardiovascular outcomes trial testing ziltivekimab will not be completed before 2025.^15^

However, any benefits from anti-IL-6 treatments would need to be balanced against an impaired host response. Canakinumab has been associated with a higher risk of fatal infections^12^ and genetic analyses support that downregulation of IL-6 signaling is indeed associated with a higher risk of bacterial infections.^11^ To this end, IL-6 is known as a key component of the host innate immune defense system^16^. As such, while trials directly targeting IL-6 signaling are ongoing, it would be useful to identify alternative drug targets in the same signaling pathway that might mediate the atheroprotective effects of IL-6 inhibition. Such molecules might be more specific and thus better anti-inflammatory treatment targets for atherosclerotic cardiovascular disease.

We previously leveraged genetic data from large-scale population-based studies and detected 26 variants in the gene encoding IL-6R that mimic the effects of pharmacologically inhibiting this protein.^17, 18^ Here, we expand these analyses to blood proteomic data derived from the INTERVAL study (n=3,301) in order to explore: (i) the causal circulating signature of interfering with IL-6 signaling that could inform the discovery of biomarker signatures of drug response to IL-6 inhibitors, and (ii) potential mediators of the favorable cardiovascular effects of IL-6 inhibition that could serve as alternative and perhaps more selective drug targets. Following-up on our findings, we compare our results to those of clinical trials applying pharmacological inhibition of IL-6 signaling. Triangulating the evidence, we explore in observational studies associations between circulating levels of potential protein drug targets and incident cardiovascular disease as well as the histological, transcriptomic, and cellular profile of atherosclerotic lesions associated with these targets.

## METHODS

### Genetic instrument selection

To construct an instrument for IL-6R-mediated signaling,^19^ we used variants within the *IL6R* gene or a region 300 kB upstream or downstream of it. In accordance with an approach that we previously described,^17, 18^ we selected variants associated in a GWAS meta-analysis of 522,681 individuals with circulating levels of C-reactive protein (CRP), a downstream biomarker of IL-6 signaling^7^ that was used as a functional readout for IL-6 signaling activity. The methodology and criteria used to obtain this instrument are outlined in the **Supplementary Methods** and the comprising genetic variants are listed in **Supplementary Table S1**. As previously described, this genetic instrument is associated with circulating IL-6, CRP, and fibrinogen levels, consistent with results of clinical trials testing tocilizumab, a monoclonal antibody against IL-6R, as has been previously described.^17^

### Proteome-wide Mendelian randomization analyses

To detect the downstream proteomic effects of genetically proxied IL-6 signaling, we applied a proteome-wide two-sample Mendelian randomization approach using plasma data from the INTERVAL study (n=3,301 with SOMAscan® aptamer-based proteomics), more information on which is available in the **Supplementary Methods**. Using these data, we applied two-sample MR analysis applying the inverse-variance weighted (IVW) method as our primary approach. As a measure of pleiotropy, heterogeneity was quantified with the I^2^ and significance was tested with the Cochran Q statistic (p<0.05 was considered significant).^20^ To correct for multiple comparisons across the proteome, we corrected the p-values derived from the IVW approach using the false discovery rate (FDR) method.^21^ P values<0.05 were considered statistically significant. In order to validate our results against bias arising from horizontal pleiotropy, for significant proteins, we also applied the weighted median approach,^22^ which is more robust to the use of pleiotropic instruments. Furthermore, for significant proteins, we applied a colocalization analysis, in order to ensure that the same genetic signal influencing IL-6 signaling also influences protein levels and that the detected association was not the result of pleiotropic effects of variants in LD with the selected instruments.^23^ We applied a Bayesian framework for pairwise colocalization, within 300 kB of the *IL6R* gene using the coloc package in R and tested competing models (H0: the genomic region does not include any variant influencing either the exposure or the outcome; H1: the genomic region includes a variant influencing only the exposure; H2: the genomic region includes a variant influencing only the outcome; H3: the genomic region includes separate variants influencing the exposure and the outcome; H4: the genomic region includes a variant that influences both the exposure and the outcome).^24^ The prior probability of any random variant being associated with both traits was set at 10^−5^. A posterior probability of association of 0.8 or higher for the last model (Η4) was defined as providing evidence of colocalization. The effects of the selected genetic variants on proteins were extracted from the PhenoScanner database through the phenoscanner function in R^25^ and MR analyses were performed in R using the TwoSampleMR package.^26^ Pooling the significant proteins, we explored enrichment in biological pathways using the Gene Ontology (GO) resource.^27^

### Comparisons of the proteomic effects of genetically proxied IL-6 signaling downregulation with the pharmacological effects of IL-6R inhibition

To test the validity of our approach for detecting the downstream proteomic signature of a signaling pathway with proteome-wide MR, we aimed to correlate the effects derived from our MR approach with the effects derived from pharmacological IL-6R inhibition. We used data from a follow-up study of The Norwegian tocilizumab non–ST-segment–elevation MI (NSTEMI) trial, which tested IL-6 blockade with tocilizumab in patients with NSTEMI. In the original randomized control trial patients with NSTEMI received a single intravenous dose of tocilizumab 280mg or placebo (n=117).^28^ In a follow-up analysis, the proteomic profile of serum samples was quantified at baseline and four days post-treatment among a subgroup of 48 patients from the tocilizumab arm using the SOMAscan® aptamer-based proteomics assay.^29^ After matching the proteins available in both datasets, we quantified the Pearson’s *r* correlation between the MR effects and the pharmacological effects across the proteome.

### Associations with clinical endpoints in mediation MR analyses

After detecting proteins significantly associated with genetically proxied IL6 signaling, we then explored associations between genetically proxied levels of these proteins and the three primary manifestations of atherosclerosis in order to identify which of these proteins could mediate the effects of IL-6 signaling on clinical endpoints. Clinical outcomes included coronary artery disease (CAD), peripheral artery disease (PAD) and large-artery atherosclerotic stroke (LAAS). For CAD, we used data from 63,746 CAD cases and 130,681 controls of predominantly European ancestry (about 80%) from the CARDIoGRAMplusC4D Consortium.^30^ For PAD, we used summary statistics from the Million Veteran Program cohort, a longitudinal cohort study containing electronic health records and genetic data across 50 Veterans facilities in the US. This cohort includes 31,307 cases and 211,753 controls of European, African and Hispanic ancestry.^31^ For LAS we leveraged data from the MEGASTROKE consortium of primarily individuals of European ancestry, consisting of 6,688 stroke cases and 245,201 controls from 29 studies.^32^

Again, we applied two-sample IVW MR analyses using as instruments genetic variants associated with the proteins that were significantly associated with genetically proxied IL-6 signaling in the previous step. To avoid overlap in the used datasets, for this step, we used data from the deCODE study in the Icelandic population.^33^ This study explored the genetic architecture of 4,907 circulating proteins quantified with the aptamer-based SOMAScan® approach among 35,559 Icelanders.^33^ For the proteins significantly associated with genetically proxied IL-6 signaling, we selected genetic variants throughout the genome associated with the levels of these proteins at a p<5−10^−8^ and clumped at an *r*^2^<0.01. For all proteins, beyond the IVW approach, we applied the MR-Egger and the weighted median methods. Furthermore, for significant proteins, we applied *cis*-MR analyses, selecting genetic variants associated with the proteins within or 300 kB upstream and downstream of the genes encoding these proteins at p<10^−5^ after clumping at r^2^<0.1. Significant results were screened based on an FDR-corrected p <0.05.

For proteins showing significant associations with both genetically proxied IL-6 signaling and risk of cardiovascular endpoints, we then applied two-step mediation MR analyses^34^ to explore whether any of the effects of IL-6 signaling on the outcomes could be explained by changes in circulating protein levels. We first performed multivariable MR exploring associations of genetic proxies for significant proteins on the risk of the cardiovascular endpoints adjusting for the effects of the respective genetic instruments on sIL6R concentration. Then, by multiplying the effects of the genetic proxies for IL-6 signaling on the protein levels with the multivariable MR association estimates between genetically proxied protein levels and the cardiovascular outcomes, we obtained the indirect effects of the genetic proxies for IL-6 signaling on the outcomes mediated through the tested protein.^34^ We divided these estimates by the total effects of the genetic proxies of IL-6 signaling on risk of CAD, PAD, and LAS and obtained the proportions of the effects mediated through the tested proteins.

### Replication in an observational population-based setting

CXCL10/interferon-γ inducing protein 10 (IP-10) showed particularly promising associations as a potential mediator of genetically proxied IL-6 signaling on all tested atherosclerotic outcomes (as shown in the Results section). To replicate these associations in an observational setting, we used data from the population-based MONICA/KORA Cohort,^35^ a prospective population-based study of inhabitants of Augsburg, Germany, aged 25-74 years at baseline (1984-1995) followed-up until 2016. Further information on the study design, outcome assessment, the characteristics of the selected study participants and the CXCL10 measurement methodology can be found in the **Supplementary Methods**.

To explore associations between baseline CXCL10 levels with risk of future cardiovascular events (stroke or CAD), we applied a Cox proportional hazards model adjusting for age, sex, baseline survey (model 1), and additionally for vascular risk factors (body mass index (1 kg/m^2^ increment), smoking (current vs. non-current), estimated glomerular filtration rate (1 mL/min/1.73 m² increment), history of CAD, diabetes mellitus, total cholesterol, HDL cholesterol, and hypertension defined as blood pressure >140/90 mmHg or use of antihypertensive medications if participants were aware of having hypertension) (model 2). Based on the assumption that CXCL10 is a downstream effector of IL-6 signaling on atherosclerosis, we explored correlations between baseline IL-6 and CXCL10 levels, and in sensitivity analyses included IL-6, CXCL10, or both in our models to see how the effects of each cytokine on cardiovascular events changed after adjusting for the other.

### Analyses in human atherosclerotic samples

We used data from the Athero-EXPRESS study, a biobank of carotid endarterectomy samples, where the expression levels of CXCL10 have been quantified in transcriptomic analyses, to assess associations with histological features of plaque vulnerability. For 700 samples, RNA was isolated and libraries were prepared for sequencing as previously described.^36^ We tested associations between the normalized expression values of CXCL10 and standard histological features of plaque vulnerability (large lipid core, intraplaque hemorrhage, extensive collagen content, and plaque calcification), as determined in sections from the plaque segment with the highest atherosclerosis burden. Methods for the evaluation of the histological images have been previously described in detail.^37^

To explore the changes in atherosclerotic plaque associated with higher expression of CXCL10, we used data from the Stockholm-Tartu Atherosclerosis Reverse Networks Engineering Task (STARNET). We used transcriptomics data from atherosclerotic aortic root samples obtained with informed consent during coronary artery bypass grafting (CABG) from 514 individuals with CAD.^38^ We explored genes co-expressed with CXCL10 by estimating Spearman correlations with the expression of 16,214 genes in this tissue. Genes significantly co-expressed with CXCL10 were moved forward to pathway analyses in Reactome.^39^ To explore the cell landscape of the transcriptomic profile associated with upregulated *CXCL10* expression, we then used publicly available single nuclei transcriptomic data from aortic tissue and tested which of the identified cell groups were enriched for these genes.^40^

### Data availability and ethical approval

Details for access to summary statistics used for the current analyses are provided in the **Supplementary Methods**. All included studies have received ethical approval from the respective institutional review boards and all participants have provided informed consent, as detailed in the **Supplementary Methods**.

## RESULTS

### Effects of genetically proxied IL-6 signaling on circulating proteins

The overall study design is presented in **Fig. 1**. The 26 CRP-lowering genetic variants within or close to the gene encoding IL-6R that were used as proxies for IL-6 signaling downregulation are presented in **Supplementary Table S1**. First, we explored associations between genetically proxied IL-6R-mediated signaling as captured by these variants, with proteomic changes across 3,281 plasma proteins quantified with the aptamer-based SOMAScan assay among 3,301 participants of the INTERVAL study, as illustrated in **Fig. 2a**. A detailed list of the associations as derived from IVW MR analyses is provided in **Supplementary Table S2**. After correction for multiple comparisons (FDR-adjusted p-value<0.05), we found that genetically proxied downregulation of the IL-6 signaling pathway is associated with lower levels of 43 and higher levels of 27 circulating proteins (**Fig. 2b** and **Supplementary Table S3**). Of those proteins, 54 also showed evidence of colocalization, as determined by a PPA>0.80 for at least one common genetic variant in the examined genomic region influencing both sIL6r and the respective protein levels. Among these proteins, 48 also showed significant and directionally consistent associations in the weighted median approach. Of those, only one showed minimal evidence of heterogeneity (MYO6, I^2^=27%, **Supplementary Table S3**).

**Figure 1.**
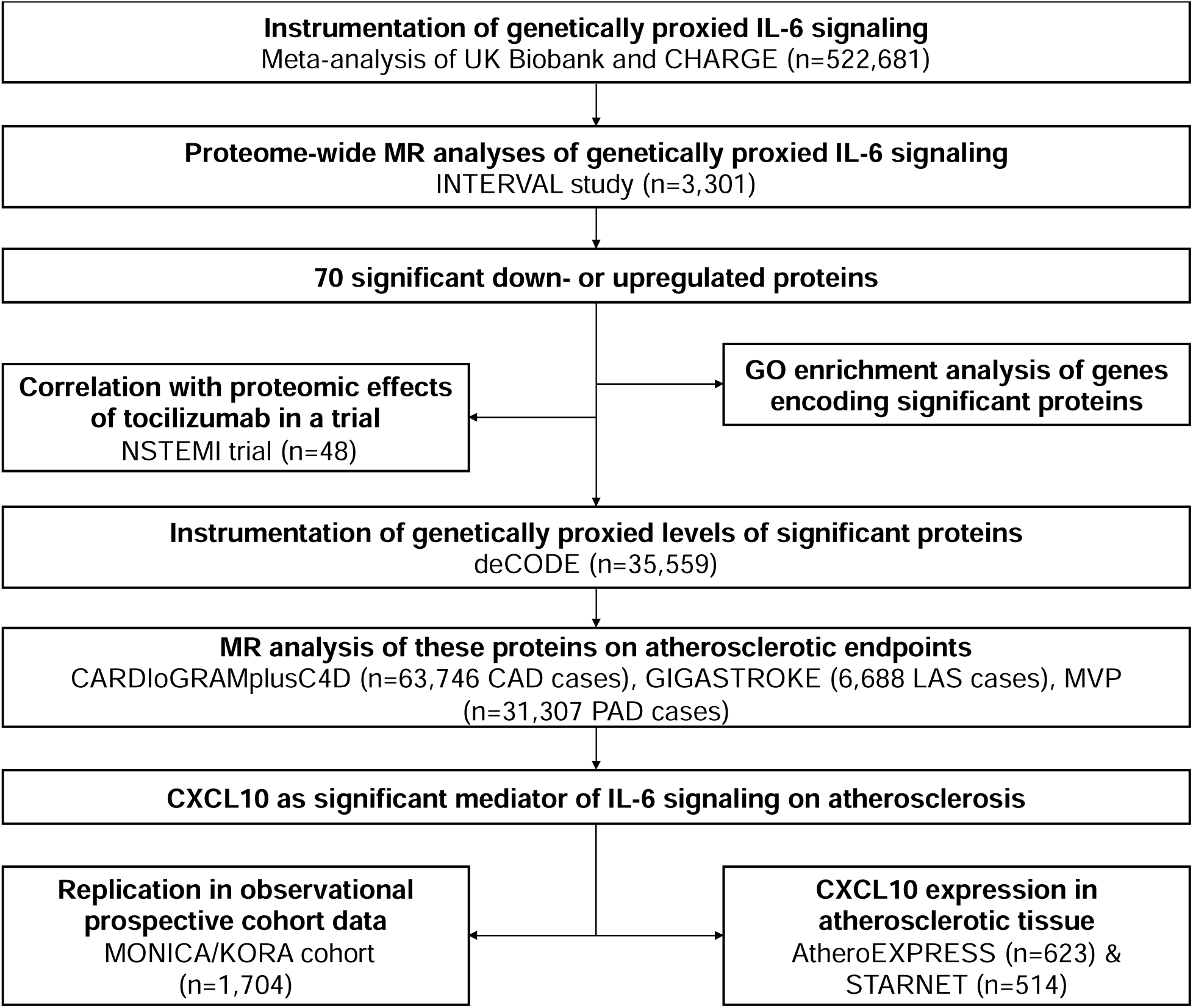
Study overview. Overview of major analytical approaches and data sources used in the current study to identify proteins up- or downregulated by genetically proxied IL-6 signaling as well as mediators of its effect on atherosclerotic disease.

**Figure 2.**
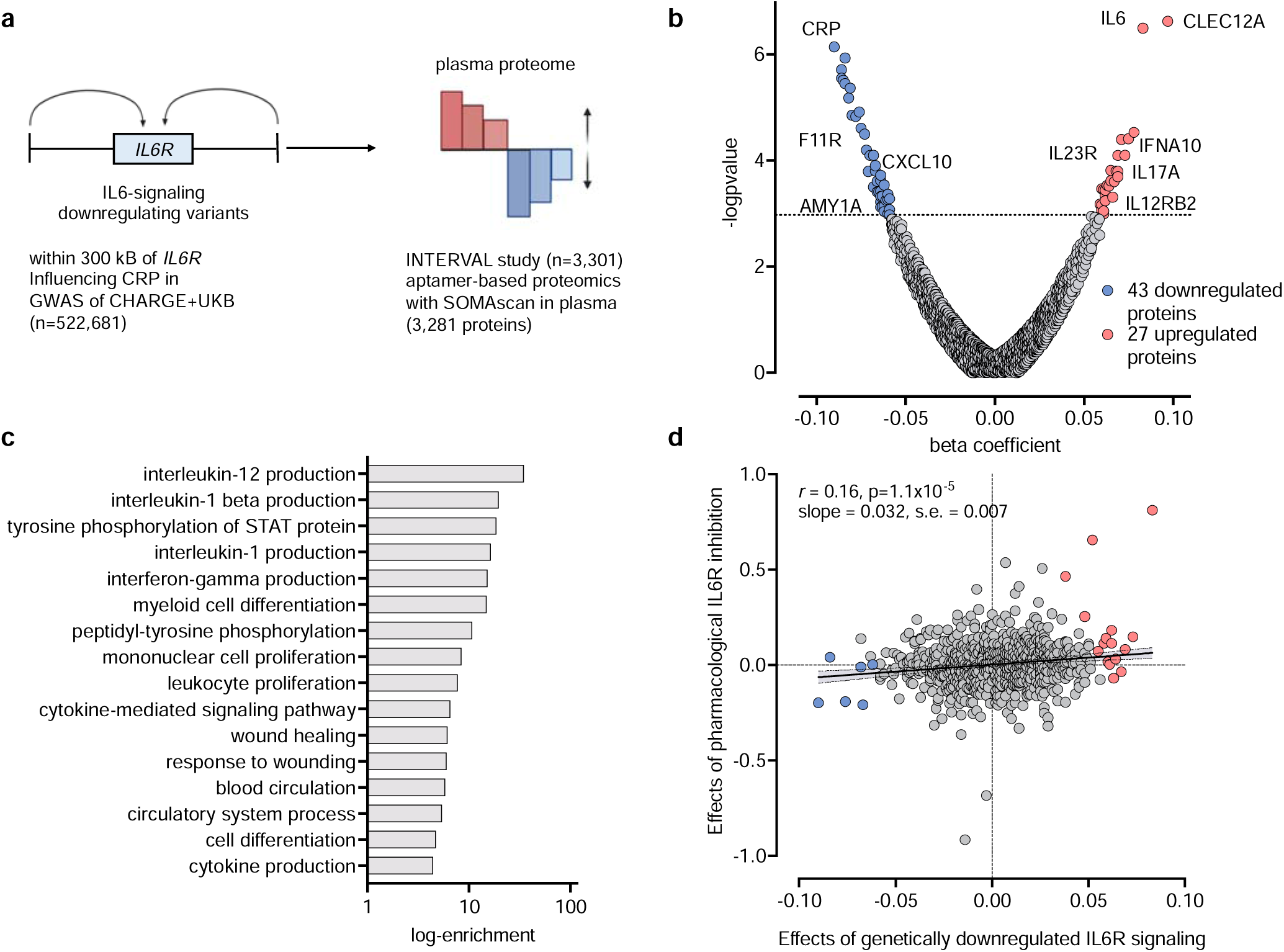
Plasma proteomic changes in association with genetically proxied IL-6 signaling. (**a**) Schematic representation of the study design and data sources. (**b**) Volcano plot of the associations of genetically downregulated IL-6 signaling with plasma proteins in the INTERVAL study (n=3,301). The results are derived from random-effects inverse-variance weighted Mendelian randomization analyses. The dotted line corresponds to a false discovery rate (FDR)-corrected p-value<0.05. (**c**) Significant (FDR-corrected p-value<0.05) Gene Ontology (GO) Pathway enrichment analysis for significant proteins. (**d**) Correlation between Mendelian randomization estimates for proteins associated with genetically downregulated signaling and estimates from linear regression for pharmacological IL-6R inhibition among 24 individuals treated with tocilizumab versus 24 individuals treated with placebo in the Norwegian tocilizumab NSTEMI study (the blue and red dots correspond to proteins that were found to be down- and upregulated, respectively in the genetic analysis of panel B).

Among the 70 proteins, there were well-known components of the pathway, such as IL-6 itself or downstream mediators and effectors, such as CRP and the fibrinogen C-domain-containing protein-1 (**Fig. 2b** and **Supplementary Table S3**). A GO pathway enrichment analysis with the significant proteins (**Fig. 2c** and **Supplementary Table S4**) showed enrichment in several cytokine production pathways, such as regulation of IL-12 and IL-1β production and regulation of the ΙFN-γ pathway, as well as in immune cell trafficking pathways, such as myeloid cell differentiation and mononuclear cell or leukocyte proliferation. Furthermore, there was enrichment in tyrosine phosphorylation of STAT in line with the known signaling transduction pathway of IL-6R activation.^41^

### Correlation between MR-derived proteomic and pharmacological effects

We compared the proteomic effects derived from a genetically proxied downregulation of IL-6 signaling in MR analyses with the effects of pharmacological IL-6 signaling inhibition on the same proteins. Leveraging data from 48 patients with NSTEMI from the Norwegian tocilizumab NSTEMI study (24 who underwent treatment with tocilizumab and 24 in the control arm),^29^ we obtained differences between tocilizumab and placebo across 1074 proteins also assessed with a different version of the aptamer-based SOMAScan assay within 48 hours post-randomization and treatment. Matching the available proteins with the proteins available in the INTERVAL study, we were able to explore correlations across 785 proteins between the MR and the trial effects (**Supplementary Table S5**). We found a significant correlation across the proteome (Pearson’s *r* 0.16, p=1.1−10^−5^, **Fig. 2d**). This correlation was even stronger when restricting the analysis to the 44 proteins reaching a p<0.05 in the trial analysis (r=0.48, p=8−10^−4^).

### Effects of significant proteins on cardiovascular endpoints and mediation analysis

Given the well-established associations of genetically downregulated IL-6 signaling with CAD, PAD, and ischemic stroke,^8, 9, 11^ we next explored to what extent the 70 proteins associated with genetically proxied IL-6 signaling are also associated with these cardiovascular endpoints. We proxied the levels of 58 of these proteins in the deCODE proteogenomic database from 35,559 Icelanders, where 4,907 circulating proteins were also quantified with the aptamer-based SOMAScan assay.^33^ Across these 58 proteins, we found the genetically proxied circulating levels of four of them to be associated with the examined cardiovascular outcomes in IVW MR analyses following correction for multiple comparisons (FDR-corrected p-value<0.05). Genetically proxied levels of CXCL10 were associated with all three atherosclerosis endpoints, whereas cystathionine β-synthase (CBS), CRP, and interleukin-23 receptor (IL-23R) showed significant associations only with CAD only (**Fig. 3a** and **Supplementary Table S6**).

**Figure 3.**
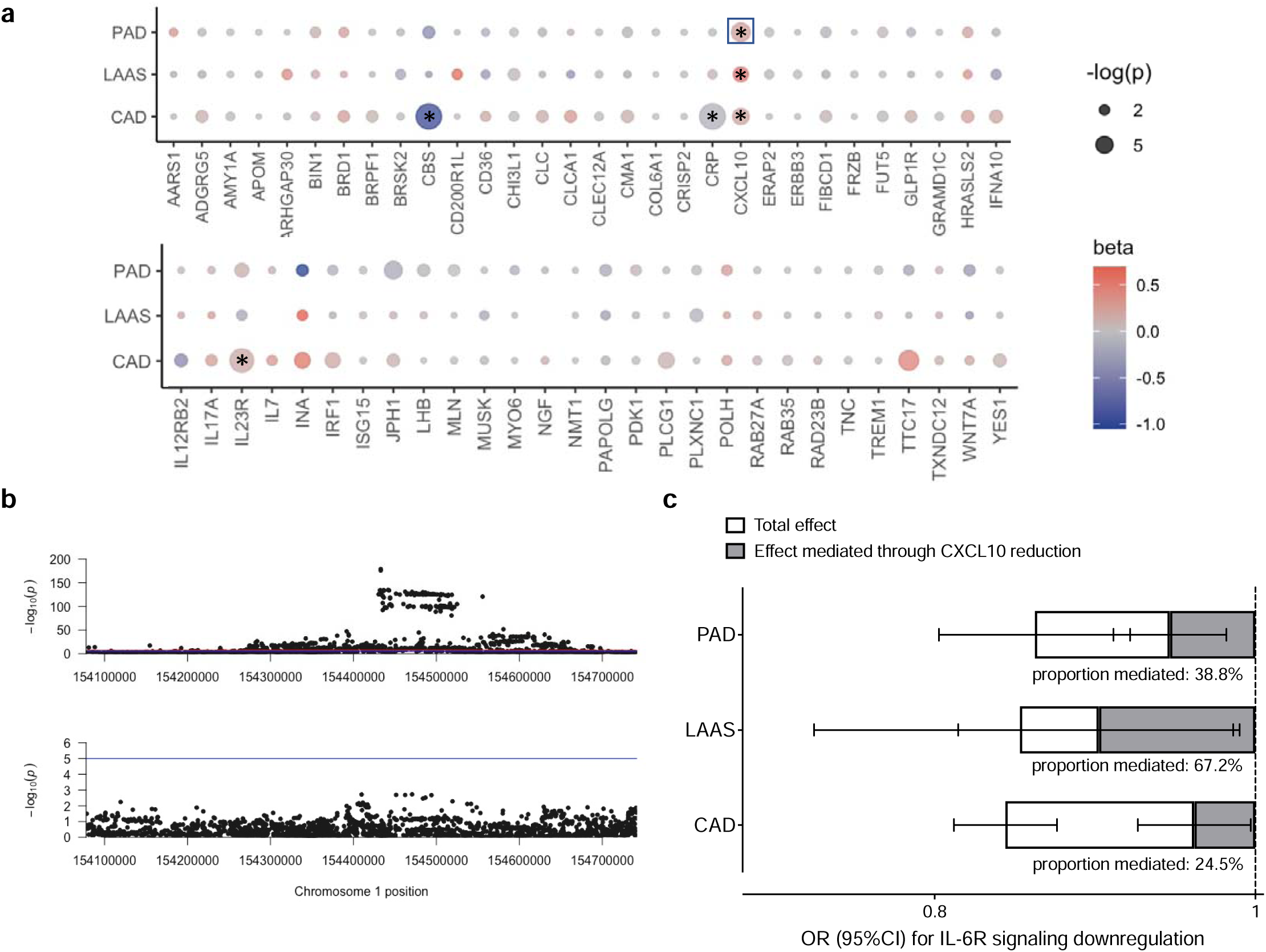
Genetically proxied levels of proteins associated with genetically downregulated IL-6 signaling and atherosclerotic cardiovascular disease. (**a**) Associations of genetically proxied levels of proteins associated with genetically downregulated IL-6 signaling with peripheral artery disease (PAD), large artery atherosclerotic stroke (LAS), and coronary artery disease (CAD), as derived from inverse-variance weighted Mendelian randomization analyses. The stars indicate significant associations at an FDR-corrected p<0.05, whereas the box around significant associations highlights associations that were also significant in cis-MR analyses. (**b**) Regional association plots at the *IL-6R* locus for associations with soluble IL-6 receptor levels (upper part) and CXCL10 levels (lower part) demonstrating colocalization of the signal. (**c**) Mediation Mendelian randomization analysis for the total effects of genetically downregulated IL-6 signaling on PAD, LAS, and CAD, as well as the indirect effects mediated through changes in CXCL10 levels.

To minimize the risk of pleiotropic effects, we next performed a sensitivity analysis based solely on *cis*-acting variants for these proteins. Using genetic variants in minimal LD (r^2^<0.1) associated with the circulating levels of these proteins at p<10^−5^ and within 300 kB upstream or downstream of the encoding genes, we confirmed the association between genetically proxied CXCL10 levels and odds for PAD (**Supplementary Table S7**). **Fig. 3b** illustrates the colocalization of the genomic signals between circulating sIL6R and CXCL10 around the same genetic variants at the *IL6R* locus. Given the emergence of CXCL10 as the most promising mediator in the examined associations, we performed mediation MR analyses. We found changes in circulating CXCL10 levels to mediate 39%, 67%, and 25% of the effects of genetically proxied IL-6 signaling on PAD, LAAS, and CAD, respectively (all p<0.05) (**Fig. 3c**).

### Circulating CXCL10 levels and risk of cardiovascular events in population-based prospective cohort study

Following-up on the signal for CXCL10, we used data from the population-based MONICA/KORA study,^35^ where circulating levels of CXCL10 have been quantified with a Luminex cytokine multiplex assay at midlife in 1,704 individuals (47% females; median age: 53 years [interquartile range: 44-61]) without overt cardiovascular disease at baseline. The baseline characteristics of the study participants are presented in **Supplementary Table S8**. There was a significant positive correlation between serum IL-6 and CXCL10 levels among study participants (r=0.19, p=1.2−10^−14^, **Fig. 4a**). Next, we tested associations of circulating CXCL10 levels with the risk of a composite endpoint of stroke and CAD over a median follow-up of 20.9 years (interquartile range: 12.9-26.1). Following adjustments for age, sex, and baseline survey (Model 1) in a Cox regression model, we found circulating CXCL10 levels to be associated with a higher risk of the composite endpoint (HR per SD increment in log-transformed CXCL10 levels: 1.20, 95%CI: 1.05-1.36, **Fig. 4b** and **4c**). These results remained significant in a model further adjusting for common vascular risk factors at baseline (Model 2), as well as when including both circulating CXCL10 and IL-6 levels in the same model (Model 3, **Fig. 4c** and **Supplementary Table S9**).

**Figure 4.**
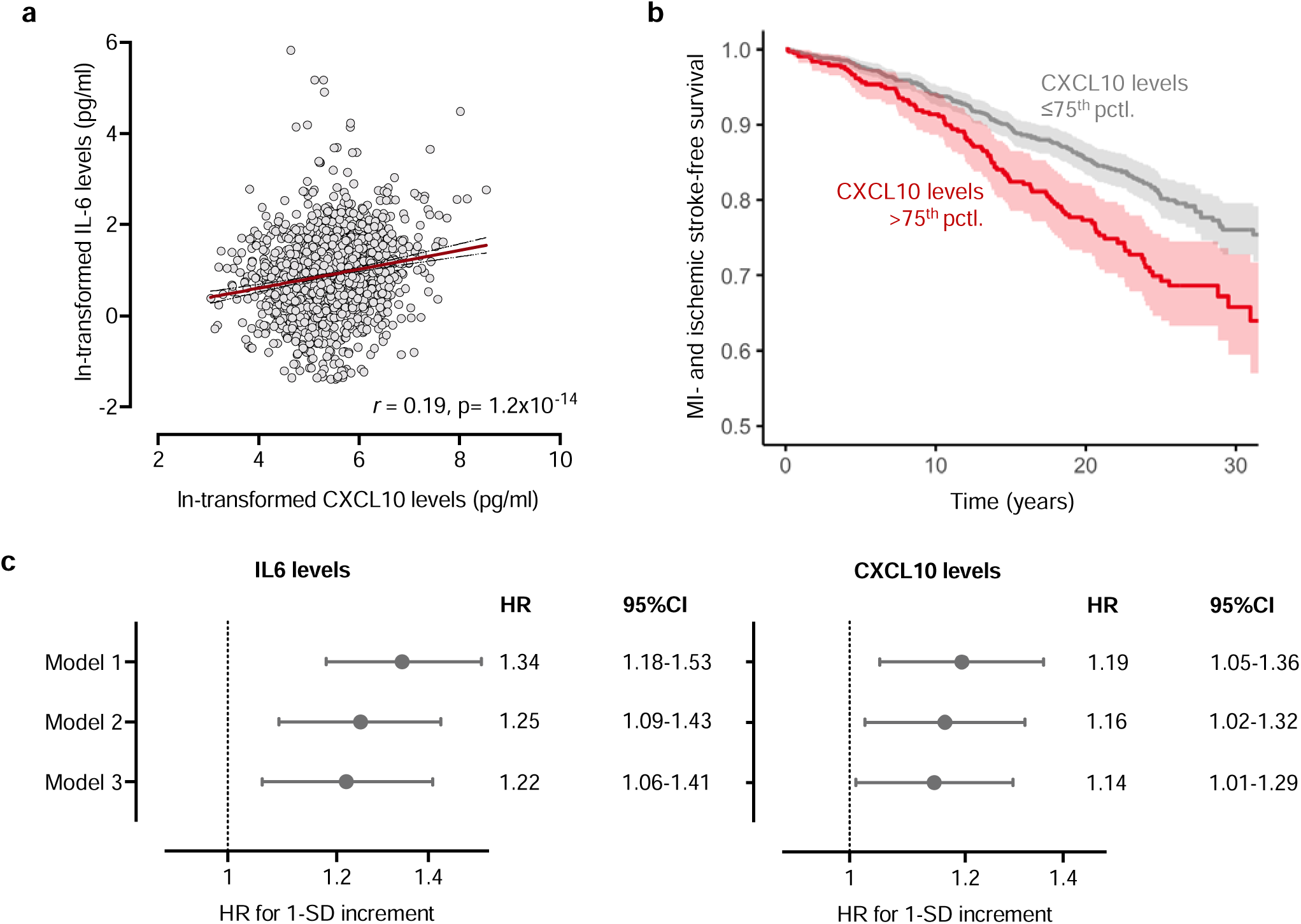
Circulating CXCL10 levels in association with circulating IL-6 and major adverse cardiovascular events in the population-based MONICA/KORA cohort. (**a**) Correlations between circulating IL-6 and CXCL10 levels among 1,704 participants of the MONICA/KORA cohort. (**b**) Kaplan-Meier curve of the associations between baseline circulating CXCL10 levels and risk of CAD or stroke over a follow-up period extending up to 30 years. (**c**) Hazard ratios of the associations of circulating IL-6 and CXCL10 levels with risk of CAD or stroke in models adjusted for age, sex, and baseline survey (Model 1), models adjusted for age, sex, baseline survey, and vascular risk factors (Model 2), and a model adjusted for all variables of Model 2 and both proteins simultaneously included (Model 3).

### CXCL10 expression in atherosclerotic tissue associated with plaque vulnerability

To explore whether CXCL10 expression in human atherosclerotic lesions is associated with plaque vulnerability, we used data from 623 individuals from the Athero-EXPRESS Biobank in Utrecht, Netherlands, who underwent carotid endarterectomy due to symptomatic or asymptomatic stenosis and for whom both transcriptomic and histological analyses were available (**Supplementary Table S10** and **Fig. 5a**). Following adjustments for age and sex, we found that higher normalized CXCL10 expression in carotid atherosclerotic plaques was associated with a larger lipid core (OR: 1.19, 95%CI: 1.02-1.40, p=0.01, **Fig. 5b**). No associations were detected with intraplaque hemorrhage, extensive collagen content, or plaque calcification (**Fig. 5b**).

**Figure 5.**
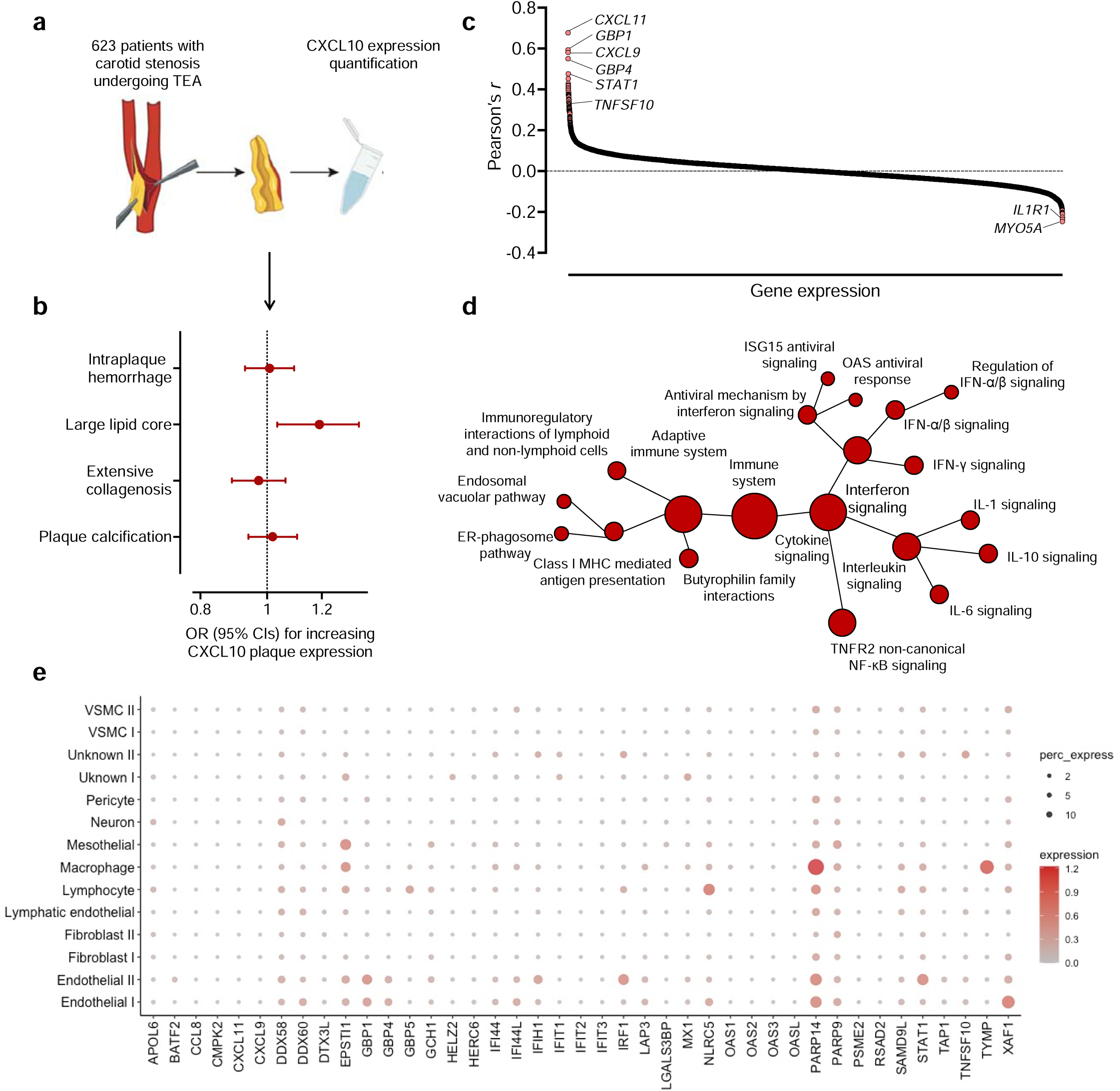
Expression of *CXCL10* within atherosclerotic lesions. (**a**) Schematic representation of CXCL10 mRNA quantification among 623 carotid atherosclerotic plaques from patients with carotid stenosis in the Athero-EXPRESS Biobank, who underwent endarterectomy. (**b**) Associations of plaque *CXCL10* expression with histological features of plaque vulnerability. (**c**) Co-expression of *CXCL10* with 16,214 other genes in atherosclerotic aortic root tissue from 514 participants in the STARNET network. (**d**) Pathway enrichment analysis of genes co-expressed with *CXCL10*, as derived from Reactome. (**e**) Cell-specific expression of top 42 genes co-expressed with CXCL10 at an *r*>0.3 in aortic tissue, as derived from single-nuclei RNA sequencing analysis in human aorta samples.

To test in more depth the phenotypic plaque profile associated with elevated *CXCL10* expression, we used transcriptomic data from atherosclerotic aortic root tissue from 514 samples from patients with CAD in the STARNET network and explored in a co-expression analysis the genes that were correlated with the expression of *CXCL10*. A total of 98 genes were positively correlated with *CXCL10* expression and 6 negatively with *CXCL10* expression (**Fig. 5c** and **Supplementary Table S11**). The positively correlated genes were enriched in immune system pathways, as captured by Reactome, and included CXCL9 and CXCL11, which are also ligands of the CXCR3, the main CXCL10 receptor.^42^ These pathways included cytokine signaling pathways, and particularly the interferon and interleukin pathways, such as IFN-γ, IL-6 and IL-1 signaling, as well as pathways related to the adaptive immune system (**Fig. 5d** and **Supplementary Table S12**). As a final step, we explored whether specific cell types, as captured by single-nuclei RNA sequencing from aortic tissue,^40^ are enriched for expressing the top genes positively co-expressed with *CXCL10* (*r*>0.3), in order to infer the cellular landscape associated with higher *CXCL10* expression. According to these data, endothelial cells, lymphocytes, and macrophages were the cell types primarily expressing these genes (*PARP14* and *STAT1* most strongly enriched in endothelial cells, *NLRC5* and *PARP14* most strongly enriched in lymphocytes, *PARP14* and *TYMP* most strongly enriched in macrophages, **Fig. 5e** and **Supplementary Table S13**).

## DISCUSSION

Integrating data from large-scale genomic and proteomic studies, we explored the downstream effects of genetic downregulation of the IL-6R-mediated signaling on the blood proteome and their mediating role in lowering the risk of atherosclerotic cardiovascular disease. We found changes in circulating levels of 70 proteins involved in regulation of cytokine production and immune cell trafficking, which were consistent with the proteomic effects of the pharmacological inhibition of IL-6R with tocilizumab in a clinical trial. Across these proteins, genetically proxied levels of the chemokine CXCL10 were associated with risk of CAD, LAAS, and PAD and changes in CXCL10 levels mediated a significant proportion of the effects of IL-6 signaling on these endpoints. In follow-up analyses, we found circulating CXCL10 levels at midlife in a population-based setting to be associated with the long-term risk of CAD or stroke. Higher expression of CXCL10 in human atherosclerotic plaques was correlated with a larger lipid core and a transcriptomic profile consistent with macrophage and lymphocyte infiltration, as well as activation of the adaptive immune system and cytokine signaling.

Our results linking CXCL10 levels with risk of atherosclerotic disease are consistent with experimental data showing a key role for this chemokine in atherogenesis and atheroprogression. The administration of a monoclonal antibody against CXCL10 produced a histologically more stable atherosclerotic plaque phenotype with increased collagen and smooth muscle cell content and a smaller necrotic lipid core in the common carotid arteries of atheroprone *Apoe^−/−^* mice fed a high-fat diet.^43^ This is in line with our results suggesting an association of higher intraplaque *CXCL10* expression with a larger lipid core, a hallmark of unstable plaques. In another model, *Cxcl10^−/−^ Apoe^−/−^* mice exhibited significantly smaller atherosclerotic lesion areas across the aortic arch, thoracic and abdominal aorta, both sexes, and different durations of treatment.^44^ The atheroprotective effect of CXCL10 blockade in translational experiments is further corroborated by evidence of decreased atherosclerotic burden in the aortic arches and carotid arteries of *Ldlr^−/−^* mice after pharmacological targeting of CXCR3, the receptor of CXCL10, with a specific small molecule antagonist^45^ and a CXCR3/CCR5 dual inhibitor.^46^ Furthermore, experimental data also suggest that IL-6 and particularly IL-6 trans-signaling via STAT3 exerts an effect on T-cell immigration via directly regulation chemokine secretion including CXCL10.^47^ CXCL10/CXCR3 interaction induces T_H_1 cell differentiation and migration into the atherosclerotic lesion where they release tumor necrosis factor-α (TNF-α) and IFN-γ thus contributing towards a proinflammatory lesion environment.^48^ Collectively, current evidence indicates that CXCL10 binding to CXCR3 drives atherosclerotic plaque progression through differentiation and homing of proinflammatory T lymphocyte subsets. More specifically, two isoforms (CXCR3-A and CXCR3-B) of this receptor have been identified in humans which lead to different downstream effects. Through binding to the CXCR3-A receptor CXCL10 has chemoattractant properties and induces the migration, proliferation and survival of leukocytes. The CXCR3B isoform of the receptor, on the other hand, has antiangiogenic properties and inhibits cell proliferation and migration while also stimulating apoptosis.^42^ It should be noted that CXCR3 also binds the chemokines CXCL9 and CXCL11.^42^

Beyond detecting mediators of the effects of IL-6 signaling on atherosclerosis, our findings also provide evidence for a downstream proteomic signature of genetic IL-6 signaling that could be correlated with trial data from pharmacological IL-6R inhibition. This finding has important implications for a more widespread use of a proteome-wide MR analysis in the context of detecting the downstream proteomic consequences of perturbing drug targets. Proteomic signatures detected with this approach could be used as potential biomarkers of response to drugs targeting the upstream targets. While such response biomarkers are well-established for drugs targeting the IL-6 signaling cascade, our approach provides proof-of-concept for the use of the method for identifying response biomarkers for drugs under earlier development. Future studies should systematically explore how proteomic alterations detected in clinical trials correlate with those detected from MR analyses.

Using MR to integrate multi-omics data is an increasingly used method for bridging data from different omics layers and detecting causal relationships between them.^49, 50^ Scaling-up mediation analyses to be integrated to this approach allows explorations of associations with clinically relevant outcomes, and as such the discovery of novel therapeutic targets. Furthermore, this approach enables the dissection of pathways leading from genomic alterations to cardiovascular disease, thus providing novel insights into pathophysiology. Expanding the focus of the approach to other omics layers beyond proteomics, to more drug targets and more outcomes could provide novel insights in future explorations.

Our study has limitations. First, despite leveraging the largest available samples, such integrating approaches are characterized by decreasing power when moving downstream across omics layers.^51^ As such, several of our analyses, especially for the outcome of LAS, might have been relatively underpowered, highlighting the need to repeat the analyses with increasing sample sizes. Second, although the proteomic analyses across all studies were done with the SOMAscan aptamer-based assay, there were considerable differences in proteins available in the INTERVAL study, deCODE, and the Norwegian tocilizumab NSTEMI trial, which led to an incomplete overlap between the studies. Comparisons with alternative proteomic platforms, like the proximity extension assay-based Olink assay should be performed in future studies. Third, the majority of individuals explored in current analyses were of European ancestry and as such our results may not be generalizable to individuals of other ancestries. Fourth, data from deCODE were derived from an Icelandic population which has substantial differences to European populations. While integration of data from deCODE with data from European datasets has been extensively used in the past providing important insights into disease pathogenesis, we cannot exclude the introduction of population stratification that could bias our MR analyses in the second step (**Fig. 2**) of our approach. Fifth, an additional caveat to our study is that we used proxies of IL-6R inhibition as genetic instruments, meaning that proteins found upstream would be upregulated as a result of positive feedback while downstream molecules would be downregulated. This could create confusion as to the directionality of the levels of these proteins in real-life clinical trials. Sixth, to proxy IL-6 signaling we used CRP levels, which increase in response to the activation of the IL-6 cascade. However, IL-6 signaling is a complex cascade with a classical component, exerted through the membrane-bound IL-6R, and a trans-signaling component exerted through the soluble form of IL-6R. The two sub-pathways exert unique actions,^16, 52^ but disentangling them with genetic instruments goes probably beyond the limits of MR. Seventh, by design, MR analyses assess the effects of lifetime downregulated IL-6 signaling, which might differ from a shorter pharmacological inhibition with IL6-R blockade.

In conclusion, integrating genomic and proteomic data, we found a proteomic signature of IL-6 signaling activation and mediators of its effect on cardiovascular disease. Our analyses suggest CXCL10 to be a potentially causal mediator for atherosclerotic endpoints in three different vascular beds and as such might serve as a promising drug target for atheroprotection that should be further explored in clinical trials.

## Data Availability

Summary statistics of the genetic analyses from the INTERVAL and deCODE studies are publicly available online at https://ega-archive.org/datasets/EGAD00001004080/files and https://www.decode.com/summarydata/. The genetic instruments used for the main analyses are provided in Supplementary Table S1 and are available from previous publications.1,8 GWAS summary statistics for coronary artery disease and large artery stroke from the CardioGRAMPLUSC4D Consortium and the MEGASTROKE Consortium are available at http://www.cardiogramplusc4d.org/data-downloads/ and https://www.megastroke.org/download.html, respectively. GWAS summary statistics for PAD have been accessed through dbGAP (pha005161.1) with an approved application by JR and CDA for the Veterans Administration (VA) Million Veteran Program (MVP) Summary Results from Omics Studies. Summary statistics from the NSTEMI trial were accessed through contact of the corresponding authors of the study and are available to interested authors through an application and a short research proposal submitted to the study. Data from the MONICA/KORA study are not publicly available because the data are subject to national data protection laws and restrictions were imposed by the ethics committee of the Bavarian Chamber of Physicians to ensure data privacy of the study participants. However, data are available on request to researchers through a project agreement from KORA (http://epi. helmholtz-muenchen.de/kora-gen/). Requests should be sent to and are subject to approval by the KORA board. The datasets from Athero-Express analyzed for the current study are available upon reasonable request and application to Athero-Express Biobank Study through a Data Transfer Agreement due to consent restrictions and local regulations. The gene expression dataset from STARNET was accessed through dbGAP with an approved application by JR and CDA (phs001203.v1.p1). Data for the single-nuclei transcriptomic analyses were accessed through the Single Cell Portal (https://singlecell.broadinstitute.org/single_cell). The YFS is a population-based prospective follow-up study on cardiovascular risk factors in Finland. It has been carried out in all five Finnish university cities with medical schools and their rural surroundings. The first cross-sectional study was conducted in 1980. Altogether 4,320 children and adolescents aged 3, 6, 9, 12, 15 and 18 years were randomly recruited from the population register of these areas to produce a representative subsample of Finnish children. Of these individuals 3,596 (83%) participated in year 1980. Since then, regular follow-up visits have been performed in 1980, 1983, 1986, 2001, 2007, 2011 and 2018/2020. Follow-up using registry data (diagnoses, medications and mortality) have been extended by 2018. The YFS has been approved by the Joint Commission on Ethics of the Turku University and the Turku University Central Hospital and has been conducted according to the guidelines of the Declaration of Helsinki, and informed consent has been obtained from all participants or their parents.

http://www.cardiogramplusc4d.org/data-downloads/

https://www.megastroke.org/download.html

http://epi.helmholtz-muenchen.de/kora-gen/

https://singlecell.broadinstitute.org/single_cell

https://www.decode.com/summarydata/

## Supplemental Material

Extended Methods

Supplementary Tables S1-S13

## Acknowledgements

We thank all participants for their long-term commitment to the KORA study, the staff for data collection and research data management and the members of the KORA Study Group (https://www.helmholtz-munich.de/en/epi/cohort/kora) who are responsible for the design and conduct of the study.

## Non-standard Abbreviations and Acronyms

CABG: Coronary artery bypass grafting
CAD: Coronary artery disease
CANTOS: Canakinumab Anti-Inflammatory Thrombosis Outcomes Study
CBS: Cystathionine Beta-Synthase
CHARGE: Cohorts for heart and aging research in genomic epidemiology
CI: Confidence interval
CRP: C-eactive protein
CXCL10: C-X-C motif chemokine ligand 10
CXCR3: C-X-C motif chemokine receptor 3
EHR: Electronic health records
FDR: False discovery rate
GO: Gene ontology
HR: Hazard ratio
IFN-γ: Interferon gamma
IL-6: Interleukin-6
IL-1: Interleukin 1
IL6-R: Interleukin-6 receptor alpha chain
IP-10: Interferon-γ-induced protein 10)
IVW: Inverse variance weighted
LAAS: Large artery atherosclerotic stroke
LD: Linkage disequilibrium
MR: Mendelian randomization
NSTEMI: non–ST-segment–elevation MI
PAD: Peripheral artery disease
SD: Standard deviation
SE: Standard error
sIL6R: Soluble IL-6R
SLE: Systemic lupus erythematosus
STARNET: Stockholm-Tartu atherosclerosis reverse networks engineering task
TNF-α: Tumor necrosis factor-α

## REFERENCES

1. Soehnlein O, Libby P. Targeting inflammation in atherosclerosis - from experimental insights to the clinic. Nat Rev Drug Discov. 2021;20:589–610. doi: 10.1038/s41573-021-00198-1

2. Ridker PM, Rane M. Interleukin-6 Signaling and Anti-Interleukin-6 Therapeutics in Cardiovascular Disease. Circ Res. 2021;128:1728–1746. doi: 10.1161/CIRCRESAHA.121.319077

3. Schuett H, Oestreich R, Waetzig GH, Annema W, Luchtefeld M, Hillmer A, Bavendiek U, von Felden J, Divchev D, Kempf T, et al. Transsignaling of interleukin-6 crucially contributes to atherosclerosis in mice. Arterioscler Thromb Vasc Biol. 2012;32:281–290. doi: 10.1161/ATVBAHA.111.229435

4. Huber SA, Sakkinen P, Conze D, Hardin N, Tracy R. Interleukin-6 exacerbates early atherosclerosis in mice. Arterioscler Thromb Vasc Biol. 1999;19:2364–2367. doi: 10.1161/01.atv.19.10.2364

5. Kaptoge S, Seshasai SR, Gao P, Freitag DF, Butterworth AS, Borglykke A, Di Angelantonio E, Gudnason V, Rumley A, Lowe GD, et al. Inflammatory cytokines and risk of coronary heart disease: new prospective study and updated meta-analysis. Eur Heart J. 2014;35:578–589. doi: 10.1093/eurheartj/eht367

6. Papadopoulos A, Palaiopanos K, Björkbacka H, Peters A, de Lemos JA, Seshadri S, Dichgans M, Georgakis MK. Circulating interleukin-6 levels and incident ischemic stroke: a systematic review and meta-analysis of population-based cohort studies. medRxiv. 2021:2021.2003.2027.21254451. doi: 10.1101/2021.03.27.21254451

7. Interleukin-6 Receptor Mendelian Randomisation Analysis C, Swerdlow DI, Holmes MV, Kuchenbaecker KB, Engmann JE, Shah T, Sofat R, Guo Y, Chung C, Peasey A, et al. The interleukin-6 receptor as a target for prevention of coronary heart disease: a mendelian randomisation analysis. Lancet. 2012;379:1214–1224. doi: 10.1016/S0140-6736(12)60110-X

8. Georgakis MK, Malik R, Gill D, Franceschini N, Sudlow CLM, Dichgans M, Invent Consortium CIWG. Interleukin-6 Signaling Effects on Ischemic Stroke and Other Cardiovascular Outcomes: A Mendelian Randomization Study. Circ Genom Precis Med. 2020;13:e002872. doi: 10.1161/CIRCGEN.119.002872

9. Levin MG, Klarin D, Georgakis MK, Lynch J, Liao KP, Voight BF, O’Donnell CJ, Chang KM, Assimes TL, Tsao PS, et al. A Missense Variant in the IL-6 Receptor and Protection from Peripheral Artery Disease. Circ Res. 2021. doi: 10.1161/CIRCRESAHA.121.319589

10. Cai T, Zhang Y, Ho YL, Link N, Sun J, Huang J, Cai TA, Damrauer S, Ahuja Y, Honerlaw J, et al. Association of Interleukin 6 Receptor Variant With Cardiovascular Disease Effects of Interleukin 6 Receptor Blocking Therapy: A Phenome-Wide Association Study. JAMA Cardiol. 2018;3:849–857. doi: 10.1001/jamacardio.2018.2287

11. Georgakis MK, Malik R, Li X, Gill D, Levin MG, Vy HMT, Judy R, Ritchie M, Verma SS, Regeneron Genetics C, et al. Genetically Downregulated Interleukin-6 Signaling Is Associated With a Favorable Cardiometabolic Profile: A Phenome-Wide Association Study. Circulation. 2021;143:1177–1180. doi: 10.1161/CIRCULATIONAHA.120.052604

12. Ridker PM, Everett BM, Thuren T, MacFadyen JG, Chang WH, Ballantyne C, Fonseca F, Nicolau J, Koenig W, Anker SD, et al. Antiinflammatory Therapy with Canakinumab for Atherosclerotic Disease. N Engl J Med. 2017;377:1119–1131. doi: 10.1056/NEJMoa1707914

13. Ridker PM, Libby P, MacFadyen JG, Thuren T, Ballantyne C, Fonseca F, Koenig W, Shimokawa H, Everett BM, Glynn RJ. Modulation of the interleukin-6 signalling pathway and incidence rates of atherosclerotic events and all-cause mortality: analyses from the Canakinumab Anti-Inflammatory Thrombosis Outcomes Study (CANTOS). Eur Heart J. 2018;39:3499–3507. doi: 10.1093/eurheartj/ehy310

14. Ridker PM, Devalaraja M, Baeres FMM, Engelmann MDM, Hovingh GK, Ivkovic M, Lo L, Kling D, Pergola P, Raj D, et al. IL-6 inhibition with ziltivekimab in patients at high atherosclerotic risk (RESCUE): a double-blind, randomised, placebo-controlled, phase 2 trial. Lancet. 2021;397:2060–2069. doi: 10.1016/S0140-6736(21)00520-1

15. Novo Nordisk A/S. ZEUS - A Research Study to Look at How Ziltivekimab Works Compared to Placebo in People With Cardiovascular Disease, Chronic Kidney Disease and Inflammation (ZEUS). 2021.

16. Rose-John S, Winthrop K, Calabrese L. The role of IL-6 in host defence against infections: immunobiology and clinical implications. Nat Rev Rheumatol. 2017;13:399–409. doi: 10.1038/nrrheum.2017.83

17. Georgakis MK, Malik R, Burgess S, Dichgans M. Additive effects of genetic IL-6 signaling downregulation and LDL-cholesterol lowering on cardiovascular disease: a 2−2 factorial Mendelian randomization analysis. Journal of the Americal Heart Association. 2021;(In Press).

18. Georgakis MK, Malik R, Richardson TG, Howson JMM, Anderson CD, Burgess S, Hovingh GK, Dichgans M, Gill D. Associations of genetically predicted IL-6 signaling with cardiovascular disease risk across population subgroups. BMC Med. 2022;20:245. doi: 10.1186/s12916-022-02446-6

19. Georgakis MK, Malik R, Gill D, Franceschini N, Sudlow CLM, Dichgans M. Interleukin-6 Signaling Effects on Ischemic Stroke and other Cardiovascular Outcomes: A Mendelian Randomization Study. Circ Genom Precis Med. 2020. doi: 10.1161/CIRCGEN.119.002872

20. Bowden J, Hemani G, Davey Smith G. Invited Commentary: Detecting Individual and Global Horizontal Pleiotropy in Mendelian Randomization-A Job for the Humble Heterogeneity Statistic? Am J Epidemiol. 2018;187:2681–2685. doi: 10.1093/aje/kwy185

21. Aggarwal S, Yadav AK. False Discovery Rate Estimation in Proteomics. Methods Mol Biol. 2016;1362:119–128. doi: 10.1007/978-1-4939-3106-4_7

22. Hartwig FP, Davey Smith G, Bowden J. Robust inference in summary data Mendelian randomization via the zero modal pleiotropy assumption. International journal of epidemiology. 2017;46:1985–1998. doi: 10.1093/ije/dyx102

23. Zuber V, Grinberg NF, Gill D, Manipur I, Slob EAW, Patel A, Wallace C, Burgess S. Combining evidence from Mendelian randomization and colocalization: Review and comparison of approaches. Am J Hum Genet. 2022;109:767–782. doi: 10.1016/j.ajhg.2022.04.001

24. Giambartolomei C, Vukcevic D, Schadt EE, Franke L, Hingorani AD, Wallace C, Plagnol V. Bayesian test for colocalisation between pairs of genetic association studies using summary statistics. PLoS Genet. 2014;10:e1004383. doi: 10.1371/journal.pgen.1004383

25. Kamat MA, Blackshaw JA, Young R, Surendran P, Burgess S, Danesh J, Butterworth AS, Staley JR. PhenoScanner V2: an expanded tool for searching human genotype-phenotype associations. Bioinformatics. 2019;35:4851–4853. doi: 10.1093/bioinformatics/btz469

26. Hemani G, Zheng J, Elsworth B, Wade KH, Haberland V, Baird D, Laurin C, Burgess S, Bowden J, Langdon R, et al. The MR-Base platform supports systematic causal inference across the human phenome. Elife. 2018;7. doi: 10.7554/eLife.34408

27. Gene Ontology C. The Gene Ontology resource: enriching a GOld mine. Nucleic Acids Res. 2021;49:D325–D334. doi: 10.1093/nar/gkaa1113

28. Kleveland O, Kunszt G, Bratlie M, Ueland T, Broch K, Holte E, Michelsen AE, Bendz B, Amundsen BH, Espevik T, et al. Effect of a single dose of the interleukin-6 receptor antagonist tocilizumab on inflammation and troponin T release in patients with non-ST-elevation myocardial infarction: a double-blind, randomized, placebo-controlled phase 2 trial. Eur Heart J. 2016;37:2406–2413. doi: 10.1093/eurheartj/ehw171

29. George MJ, Kleveland O, Garcia-Hernandez J, Palmen J, Lovering R, Wiseth R, Aukrust P, Engmann J, Damas JK, Hingorani AD, et al. Novel Insights Into the Effects of Interleukin 6 Antagonism in Non-ST-Segment-Elevation Myocardial Infarction Employing the SOMAscan Proteomics Platform. J Am Heart Assoc. 2020;9:e015628. doi: 10.1161/JAHA.119.015628

30. Nikpay M, Goel A, Won HH, Hall LM, Willenborg C, Kanoni S, Saleheen D, Kyriakou T, Nelson CP, Hopewell JC, et al. A comprehensive 1,000 Genomes-based genome-wide association meta-analysis of coronary artery disease. Nat Genet. 2015;47:1121–1130. doi: 10.1038/ng.3396

31. Klarin D, Lynch J, Aragam K, Chaffin M, Assimes TL, Huang J, Lee KM, Shao Q, Huffman JE, Natarajan P, et al. Genome-wide association study of peripheral artery disease in the Million Veteran Program. Nat Med. 2019;25:1274–1279. doi: 10.1038/s41591-019-0492-5

32. Malik R, Chauhan G, Traylor M, Sargurupremraj M, Okada Y, Mishra A, Rutten-Jacobs L, Giese AK, van der Laan SW, Gretarsdottir S, et al. Multiancestry genome-wide association study of 520,000 subjects identifies 32 loci associated with stroke and stroke subtypes. Nat Genet. 2018;50:524–537. doi: 10.1038/s41588-018-0058-3

33. Ferkingstad E, Sulem P, Atlason BA, Sveinbjornsson G, Magnusson MI, Styrmisdottir EL, Gunnarsdottir K, Helgason A, Oddsson A, Halldorsson BV, et al. Large-scale integration of the plasma proteome with genetics and disease. Nat Genet. 2021;53:1712–1721. doi: 10.1038/s41588-021-00978-w

34. Carter AR, Sanderson E, Hammerton G, Richmond RC, Davey Smith G, Heron J, Taylor AE, Davies NM, Howe LD. Mendelian randomisation for mediation analysis: current methods and challenges for implementation. Eur J Epidemiol. 2021;36:465–478. doi: 10.1007/s10654-021-00757-1

35. Herder C, Baumert J, Thorand B, Martin S, Lowel H, Kolb H, Koenig W. Chemokines and incident coronary heart disease: results from the MONICA/KORA Augsburg case-cohort study, 1984-2002. Arterioscler Thromb Vasc Biol. 2006;26:2147–2152. doi: 10.1161/01.ATV.0000235691.84430.86

36. Mokry M, Boltjes A, Cui K, Slenders L, Mekke JM, Depuydt MAC, Timmerman N, Waissi F, Verwer MC, Turner AW, et al. Transcriptomic-based clustering of advanced atherosclerotic plaques identifies subgroups of plaques with differential underlying biology that associate with clinical presentation. medRxiv. 2021:2021.2011.2025.21266855. doi: 10.1101/2021.11.25.21266855

37. Georgakis MK, van der Laan SW, Asare Y, Mekke JM, Haitjema S, Schoneveld AH, de Jager SCA, Nurmohamed NS, Kroon J, Stroes ESG, et al. Monocyte-Chemoattractant Protein-1 Levels in Human Atherosclerotic Lesions Associate With Plaque Vulnerability. Arterioscler Thromb Vasc Biol. 2021;41:2038–2048. doi: 10.1161/ATVBAHA.121.316091

38. Franzen O, Ermel R, Cohain A, Akers NK, Di Narzo A, Talukdar HA, Foroughi-Asl H, Giambartolomei C, Fullard JF, Sukhavasi K, et al. Cardiometabolic risk loci share downstream cis- and trans-gene regulation across tissues and diseases. Science. 2016;353:827–830. doi: 10.1126/science.aad6970

39. Fabregat A, Sidiropoulos K, Viteri G, Forner O, Marin-Garcia P, Arnau V, D’Eustachio P, Stein L, Hermjakob H. Reactome pathway analysis: a high-performance in-memory approach. BMC Bioinformatics. 2017;18:142. doi: 10.1186/s12859-017-1559-2

40. Pirruccello JP, Chaffin MD, Chou EL, Fleming SJ, Lin H, Nekoui M, Khurshid S, Friedman SF, Bick AG, Arduini A, et al. Deep learning enables genetic analysis of the human thoracic aorta. Nat Genet. 2022;54:40–51. doi: 10.1038/s41588-021-00962-4

41. Heinrich PC, Behrmann I, Haan S, Hermanns HM, Muller-Newen G, Schaper F. Principles of interleukin (IL)-6-type cytokine signalling and its regulation. Biochem J. 2003;374:1–20. doi: 10.1042/BJ20030407

42. van den Borne P, Quax PH, Hoefer IE, Pasterkamp G. The multifaceted functions of CXCL10 in cardiovascular disease. Biomed Res Int. 2014;2014:893106. doi: 10.1155/2014/893106

43. Segers D, Lipton JA, Leenen PJM, Cheng C, Tempel D, Pasterkamp G, Moll FL, de Crom R, Krams R. Atherosclerotic Plaque Stability Is Affected by the Chemokine CXCL10 in Both Mice and Humans. International Journal of Inflammation. 2011;2011:936109. doi: 10.4061/2011/936109

44. Heller EA, Liu E, Tager AM, Yuan Q, Lin AY, Ahluwalia N, Jones K, Koehn SL, Lok VM, Aikawa E, et al. Chemokine CXCL10 Promotes Atherogenesis by Modulating the Local Balance of Effector and Regulatory T Cells. Circulation. 2006;113:2301–2312. doi: 10.1161/CIRCULATIONAHA.105.605121

45. van Wanrooij EJA, de Jager SCA, van Es T, de Vos P, Birch HL, Owen DA, Watson RJ, Biessen EAL, Chapman GA, van Berkel TJC, et al. CXCR3 Antagonist NBI-74330 Attenuates Atherosclerotic Plaque Formation in LDL Receptor–Deficient Mice. Arteriosclerosis, Thrombosis, and Vascular Biology. 2008;28:251–257. doi: 10.1161/ATVBAHA.107.147827

46. van Wanrooij EJA, Happé H, Hauer AD, de Vos P, Imanishi T, Fujiwara H, van Berkel TJC, Kuiper J. HIV Entry Inhibitor TAK-779 Attenuates Atherogenesis in Low-Density Lipoprotein Receptor–Deficient Mice. Arteriosclerosis, Thrombosis, and Vascular Biology. 2005;25:2642–2647. doi: 10.1161/01.ATV.0000192018.90021.c0

47. McLoughlin RM, Jenkins BJ, Grail D, Williams AS, Fielding CA, Parker CR, Ernst M, Topley N, Jones SA. IL-6 trans-signaling via STAT3 directs T cell infiltration in acute inflammation. Proc Natl Acad Sci U S A. 2005;102:9589–9594. doi: 10.1073/pnas.0501794102

48. Szentes V, Gazdag M, Szokodi I, Dézsi CA. The Role of CXCR3 and Associated Chemokines in the Development of Atherosclerosis and During Myocardial Infarction. 2018;9. doi: 10.3389/fimmu.2018.01932

49. Haglund A, Zuber V, Yang Y, Abouzeid M, Feleke R, Ko JH, Nott A, Babtie AC, Mills JD, Muhammed L, et al. Single-cell Mendelian randomisation identifies cell-type specific genetic effects on human brain disease and behaviour. bioRxiv. 2022:2022.2011.2028.517913. doi: 10.1101/2022.11.28.517913

50. Zheng J, Haberland V, Baird D, Walker V, Haycock PC, Hurle MR, Gutteridge A, Erola P, Liu Y, Luo S, et al. Phenome-wide Mendelian randomization mapping the influence of the plasma proteome on complex diseases. Nat Genet. 2020;52:1122–1131. doi: 10.1038/s41588-020-0682-6

51. Sadler MC, Auwerx C, Lepik K, Porcu E, Kutalik Z. Quantifying the role of transcript levels in mediating DNA methylation effects on complex traits and diseases. Nat Commun. 2022;13:7559. doi: 10.1038/s41467-022-35196-3

52. Lissilaa R, Buatois V, Magistrelli G, Williams AS, Jones GW, Herren S, Shang L, Malinge P, Guilhot F, Chatel L, et al. Although IL-6 trans-signaling is sufficient to drive local immune responses, classical IL-6 signaling is obligate for the induction of T cell-mediated autoimmunity. J Immunol. 2010;185:5512–5521. doi: 10.4049/jimmunol.1002015

