## Supplemental Material for "Proteogenomic integration reveals CXCL10 as a potentially downstream causal mediator for IL-6 signaling on atherosclerosis"

Prapiadou *et al***. Dissecting the effects of IL-6 signaling on atherosclerosis: a proteome-wide Mendelian randomization study**

**1. Extended Methods**

**1.1. Genetic Instrument Selection**

**1.2. INTERVAL Study**

**1.3. MONICA/KORA Study**

**1.4. Data Availability**

**1.5. Ethical Considerations**

**1.6. References**

**Supplementary Tables S1-S13**

**1. Extended Methods**

**1.1. Genetic Instrument Selection**

As previously described,^1^ we meta-analyzed a GWAS for CRP levels of 204,402 individuals of European ancestry from the Cohorts for Heart and Aging Research in Genomic Epidemiology  (CHARGE) Consortium^2^ with data from a GWAS for CRP levels for 318,279 White British individuals in the UK Biobank dataset.^3^ We selected as instruments genetic variants associated with CRP levels at a p<5x10^-8^ after clumping for linkage disequilibrium at *r*^2^<0.1 with the 1000G European reference panel. We weighed the variants according to their effects on circulating levels of soluble IL-6R (sIL6R). We selected this approach over weighing the variants according to their effects on CRP in order to minimize the distance between genetic variation and its biological downstream effects. We obtained the effects of the variants on sIL6R levels from the INTERVAL cohort of 3,301 individuals of European ancestry.^4^

**1.2. INTERVAL Study**

The INTERVAL study is comprised of 50,000 blood donors from 25 centers across England recruited into a randomized trial of blood donation frequency. Focusing on 3,301 randomly selected trial participants of European ancestry, 3,281 proteins were quantified in plasma samples with an aptamer-based multiplex protein assay (SOMAscan®) and their genetic architecture was explored.^4^

**1.3. MONICA/KORA Study**

CXCL10 was measured in serum obtained at the baseline visit with the Luminex® multiplex technology using a Luminex 100 analyzer (Luminex Corporation, Austin, TX, recombinant proteins and antibodies purchased from R&D systems) in non-fasting samples stored at -80°C in a case-cohort study on incident coronary artery disease^5^ and type 2 diabetes^6^. For the present analysis, CXCL10 measurements within the random sub cohort of the case-cohort study were used and the follow-up period was extended up to 2016. Medical, demographic, and biochemical data were collected at baseline visits in 1984/85, 1989/90 and 1994/95. Vital status was ascertained through the population registries inside and outside the study area. Death certificates were obtained from the local health authorities. Follow-up questionnaires were sent to all participants with consent and current address information who were still alive in 1997-1998, in 2001-2002, in 2008-2009 and in 2016 to obtain information on the occurrence of chronic diseases and risk factors.

The present analysis used a combined endpoint of incident coronary artery disease and incident stroke. All coronary artery disease events occurring until the age of 74 years (or the age of 84 years from 2009 onwards) were ascertained through the local MI registry. Coronary artery disease events occurring in participants > 74 years (or > 84 years from 2009 onwards) or in participants residing outside the study area, as well as non-fatal strokes were assessed by postal follow-up questionnaires sent out to the participants at four time points during the follow-up period as described above. All self-reported incident strokes (ischemic and hemorrhagic) as well as fatal strokes identified by death certificates and self-reported MIs and death certificate-identified fatal coronary artery disease events occurring outside the study area or in persons > 74 years (or >84 years from 2009 onwards) as well as the date of diagnosis were validated using information obtained from participants' hospital records or from their attending physicians^7^.

**1.5. Ethical Considerations**

All studies have received ethical approvals from the respective ethical authorities. All participants who donated samples to deCODE gave informed consent, and the National Bioethics Committee of Iceland approved the study, which was conducted in agreement with conditions issued by the Data Protection Authority of Iceland (VSN_14-015). Personal identities for the participant’s data and biological samples were encrypted by a third-party system (Identity Protection System), approved and monitored by the Data Protection Authority. The study protocol of Athero-EXPRESS conforms to the Declaration of Helsinki and was approved by the ethics committee on research on humans of the University Medical Center Utrecht. All participants provided written informed consent. For all KORA studies as well as the MONICA/KORA case-cohort study approval has been obtained from the Ethics Committee of the Bavarian Medical Association (Bayerische Landesärztekammer). All KORA studies have been approved by the Bavarian commissioner for data protection and privacy (Bayerischer Datenschutzbeauftragter). All study participants have provided written consent after being informed about the study. All subjects have the option to restrict their consent to specific procedures, e. g. by denying storage of biosamples. All participants in INTERVAL also gave informed consent before joining the study and the study received approval from the National Research Ethics Service approved this study (11/EE/0538). Finally, the Norwegian tocilizumab NSTEMI trial has received approval from the Regional Committee for Medical and Health Research Ethics of South‐Eastern Norway and the Norwegian Medicines Agency, and conducted according to the Declaration of Helsinki. All participants provided written informed consent. The STARNET protocol has been approved from an institutional review committee (Ethics Review Committee on Human Research of the University of Tartu).

**1.6. References**

1.	Georgakis MK, Malik R, Burgess S, Dichgans M. Additive effects of genetic IL-6 signaling downregulation and LDL-cholesterol lowering on cardiovascular disease: a 2x2 factorial Mendelian randomization analysis. *Journal of the Americal Heart Association*. 2021;(In Press).

2.	Ligthart S, Vaez A, Vosa U, Stathopoulou MG, de Vries PS, Prins BP, Van der Most PJ, Tanaka T, Naderi E, Rose LM, et al. Genome Analyses of >200,000 Individuals Identify 58 Loci for Chronic Inflammation and Highlight Pathways that Link Inflammation and Complex Disorders. *Am J Hum Genet*. 2018;103:691-706. doi: 10.1016/j.ajhg.2018.09.009

3.	Sinnott-Armstrong N, Tanigawa Y, Amar D, Mars NJ, Aguirre M, Venkataraman GR, Wainberg M, Ollila HM, Pirruccello JP, Qian J, et al. Genetics of 38 blood and urine biomarkers in the UK Biobank. *bioRxiv*. 2019:660506. doi: 10.1101/660506

4.	Sun BB, Maranville JC, Peters JE, Stacey D, Staley JR, Blackshaw J, Burgess S, Jiang T, Paige E, Surendran P, et al. Genomic atlas of the human plasma proteome. *Nature*. 2018;558:73-79. doi: 10.1038/s41586-018-0175-2

5.	Herder C, Baumert J, Thorand B, Martin S, Lowel H, Kolb H, Koenig W. Chemokines and incident coronary heart disease: results from the MONICA/KORA Augsburg case-cohort study, 1984-2002. *Arterioscler Thromb Vasc Biol*. 2006;26:2147-2152. doi: 10.1161/01.ATV.0000235691.84430.86

6.	Thorand B, Kolb H, Baumert J, Koenig W, Chambless L, Meisinger C, Illig T, Martin S, Herder C. Elevated levels of interleukin-18 predict the development of type 2 diabetes: results from the MONICA/KORA Augsburg Study, 1984-2002. *Diabetes*. 2005;54:2932-2938. doi: 10.2337/diabetes.54.10.2932

7.	Then C, Then HL, Lechner A, Thorand B, Meisinger C, Heier M, Peters A, Koenig W, Rathmann W, Scherberich J, et al. Serum uromodulin and risk for cardiovascular morbidity and mortality in the community-based KORA F4 study. *Atherosclerosis*. 2020;297:1-7. doi: 10.1016/j.atherosclerosis.2020.01.030

8.	Georgakis MK, Malik R, Richardson TG, Howson JMM, Anderson CD, Burgess S, Hovingh GK, Dichgans M, Gill D. Associations of genetically predicted IL-6 signaling with cardiovascular disease risk across population subgroups. *BMC Med*. 2022;20:245. doi: 10.1186/s12916-022-02446-6
